# ADAPTATION AND FEASIBILITY OF A BRIEF, INTEGRATED COGNITIVE CONTROL TRAINING INTERVENTION FOR DEPRESSION: A PROOF-OF-CONCEPT TRIAL

**DOI:** 10.64898/2026.08.12.26360264

**Authors:** Preeti Kodancha, Himani Kashyap, Geetha Desai

## Abstract

Cognitive deficits in depression often persist despite pharmacological and psychotherapeutic treatment. Existing cognitive retraining programs are typically time- and resource-intensive, and place limited emphasis on addressing subjectively perceived cognitive difficulties or generalization of gains. This proof-of-concept study aimed to adapt the Integrated Cognitive Control Training (ICCT) into a brief format for patients with depression and to generate preliminary evidence of feasibility and effectiveness. The intervention was adapted into a manualized five-session program through a literature review, expert surveys involving clinicians and individuals with lived experience of depression, and a trial run. The study followed a single-group, open-label pre–post design (N = 16). Significant improvements were observed in cognitive flexibility (Color Trails Test-2: t = 3.52, p = 0.003, d = 0.88), depression severity (Montgomery–Åsberg Depression Rating Scale: t = 6.66, p < 0.001, d = 1.67), and subjective cognition (Perceived Deficits Questionnaire: t = 5.06, p < 0.001, d = 1.3). The intervention demonstrated high acceptability and demand. These findings suggest that the Brief ICCT is a feasible and potentially effective approach for addressing cognitive deficits, with improvements extending to depressive symptom severity and socio-occupational functioning. These proof-of-concept findings justify further evaluation of Brief ICCT in adequately powered randomized controlled trials.

## Introduction

Depression, a disorder with a lifetime prevalence reaching up to 20% worldwide, is one of the most common and debilitating mental disorders, and a major contributor to the global disease burden (Vos et al., 2020; WHO, 2021; Chen et al., 2025). Cognitive deficits are a well-documented symptom cluster associated with depressive disorders (Ravnkilde et al., 2002; Trivedi & Greer, 2014; Rock et al., 2014). Across studies, there is a definitive link between depression and deficits in learning and memory, executive function, processing speed, and attention and concentration (Pan et al., 2018). Authors even suggest that these deficits intrinsically influence risk of relapse (Majer et al., 2004; Gonda et al., 2015), as well as socio-occupational functioning and quality of life (QoL; Stordal et al., 2009; Godard et al., 2013; Evans et al., 2014; Cambridge et al., 2018). These deficits often persist in inter-episodic periods and during remission (Conradi, Ormel, & Jonge, 2010; Yamamoto & Shimada, 2012). Further, distinction exists between subjectively perceived and objectively assessed cognitive deficits— while individuals with depression often report profound impairments, neuropsychological testing typically reveals processing inefficiencies (Lahr, Beblo, & Hartje, 2007). Notably, subjective deficits more strongly predict poorer psychosocial functioning (Mowla et al., 2007).

Despite the significant impact and persisting nature of cognitive deficits in depression, standard evidence-based treatments – pharmacological (Fehnel et al., 2013; Shilyansky et al., 2016) and psychotherapeutic (Porter et al., 2015) – fail to adequately address these impairments. Cognitive training interventions offer a promising approach to address this treatment gap. Defined as structured, scalable learning activities designed to improve cognitive or socio-affective functioning, cognitive training aims to target underlying neural mechanisms of behavioral impairment to bring about clinical change (Keshavan et al., 2014). While existing evidence on cognitive training interventions in depression shows promising results in terms of the impact on various cognitive domains (Porter et al., 2013; Motter et al., 2016; Therond et al., 2021; Legemaat et al., 2022), there are several limitations. Interventions have generally been found to fall short in generalizing treatment gains to daily life (Woolf et al., 2021; Goldberg, Kuslak, & Kurtz, 2023). Existing interventions are generally time- and labor-intensive. Finally, existing interventions also do not generally target the subjectively perceived deficits in cognition. These factors likely affect the feasibility and acceptability of existing cognitive training programs and present major barriers to their widespread implementation in resource-limited low- and middle-income (LAMI) countries. As a result, cognitive training interventions are often excluded from routine clinical care in overburdened mental healthcare systems. To bridge this gap, particularly in resource-limited settings, there is a pressing need for brief, scalable, and transdiagnostic cognitive training interventions that are resource-efficient and can be delivered through technology-enabled platforms. Such approaches would enhance feasibility, improve access, and reduce the treatment gap in overburdened mental health systems. The Integrated Cognitive Control Training (ICCT) is one such promising intervention.

Originally developed for OCD, ICCT targets cognitive control and goal-directed flexibility, incorporating Bowie et al.’s (2015) ‘Pillars of Cognitive Training.’ Studies (Kashyap et al., 2019; Rini et al., 2023; Bhattacharya et al., 2023) support its efficacy in improving cognitive and generalization to socio-occupational function. Given its transdiagnostic approach and its focus on metacognitive skills grounded in evidence-based recommendations (Slagter, Davidson, & Lutz, 2011; Cicerone et al., 2019), ICCT is a suitable candidate to address the existing lacunae in cognitive training interventions for depression. However, since cognitive deficits in depression are typically less severe than those observed in OCD (Rampacher et al., 2010), the original ICCT format may not be necessary in its entirety, and a shorter, adapted version may be sufficient and more efficient for this population. Additionally, individuals with depression often report unique challenges, such as prominent subjective cognitive impairments that require targeted attention. Therefore, adapting the ICCT into a brief format targeted to the unique concerns faced by patients with depression would enhance its scalability and feasibility for use in routine clinical settings with patients with depression.

Hence, this proof-of-concept study aimed to adapt the ICCT into a brief format to address objectively-assessed and subjectively-perceived cognitive deficits in depression, and to gather preliminary evidence regarding the effectiveness and feasibility of the intervention in a tertiary healthcare setting in India.

## Materials and Methods

### Design

The study consisted of two phases.

(1) Adaptation of the ICCT to a brief format for depression.
(2) Testing the feasibility of the brief ICCT in patients with depression.

The first phase involved a comprehensive literature review and an online expert survey of clinicians and patients to identify key target areas for the intervention. The gathered data informed the adaptation of the ICCT into a brief format for patients with depression. The second phase was designed as an open-label, single-group, pre-/post-experimental design to evaluate the feasibility and efficacy of a brief version of the ICCT. Participants for the second phase were recruited between October 2023 and February 2024.

### Sample

Sample size was estimated based on the primary outcome measure (Perceived Deficits Questionnaire – Depression; PDQ-D), assuming a minimum detectable difference of 10 points with a standard deviation of 15, yielding a required sample of 24 participants for 80% power at a 5% significance level. The estimated sample size could not be achieved due to time and logistical constraints, as well as a high proportion of eligible individuals declining participation.

The final sample consisted of 16 individuals currently diagnosed with a depressive disorder. Diagnoses were made by psychiatrists, clinical psychologists, or psychiatric social workers in training, and subsequently confirmed by a licensed, consultant psychiatrist. Participants aged 18–55 years with at least a 7^th^-grade education, fluent in English or local languages (Hindi / Kannada), diagnosed with a depressive episode or dysthymia (ICD-11), and stable on medications were included. Exclusion criteria ruled out individuals with psychotic or catatonic symptoms, serious self-harm risk, intellectual disability, substance dependence (except nicotine), neurological or severe psychiatric conditions, sensorimotor impairments, recent psychological interventions (over three sessions in the past 3 months), or brain stimulation in the past three months.

The sample for this study was recruited from the outpatient psychiatry services of a tertiary psychiatric institute in India. Participants were either referred to the study by their treating clinicians or approached while on waitlists for other treatments.

### Tools

Various tools were used under the following categories: measures of subjective cognitive deficit, measures of cognitive profile with alternate forms for post-test, measures of clinical profile, measures of feasibility, and covariates, as shown in Table 1.

**Table 1.** Summary of Tools Used.

| Category | Tools |
| --- | --- |
| Subjective cognitive deficit | • Perceived Deficits Questionnaire (Sullivan et. al., 1990) |
| Metacognition | • Metacognitive Awareness and Regulation Scale (Vishwanathan et al., 2022) |
| Neuropsychological profile | • Colour Trails Test I and II (Rao et al., 2004)<br>• Digit Span (Gurappa, 2009)<br>• Spatial Span (Gurappa, 2009)<br>• Controlled Oral Word Association Test (Rao et al., 2004)<br>• Logical Memory Test (Gurappa, 2009)<br>• Block Design Test (Wechsler, 2011)<br>• Stop Signal Task (Verbruggen, Logan & Stevens, 2008) |
| Clinical profile | • Montgomery-Asberg Depression Rating Scale (Montgomery & Asberg, 1979)<br>• Hamilton Anxiety Rating Scale (Hamilton, 1959)<br>• Personality Inventory for DSM-5—Brief Form Plus Modified (Kerber et al., 2020)<br>• Clinical Global Impression – Severity Scale (Guy W., 1976) |
| Socio-occupational functioning | • Work and Social Adjustment (Mundt et al., 2002) |
| Feasibility | • Researcher-developed Checklists |
| Covariates | • Sociodemographic and clinical data sheets |

The primary outcome measures in the study are the Perceived Deficits Questionnaire (PDQ; Sullivan et. al.,1990), which measures subjectively perceived cognitive deficits, and the Color Trails Test – 2 (Christopher & Harvey, 2006), which is a measure of cognitive control.

### Intervention

The five-session intervention was conducted weekly, with each session lasting 60–90 minutes, supplemented by two weekly homework sessions of up to 20 minutes each. A session-wise manual was developed to standardize the intervention across participants, ensuring consistent delivery of all its components. The manual included detailed scripts for each session, specifying key probes, educational content, cognitive strategies, and examples to illustrate concepts effectively. Each session had a specific, structured agenda. The general structure of the sessions was as follows – (1) review of the material from the previous session and homework task; clarification of queries; (2) agenda setting for the session; (3) discussion of everyday difficulties faced in the given cognitive domain; (4) metacognitive strategy training to address the difficulties faced; (5) discussion regarding the application of strategies to everyday life; (6) in-session tasks; (7) setting homework tasks for the next session; (8) summarizing of session. An intervention fidelity checklist was also maintained to ensure that the researcher adhered to the treatment manual, and any additional techniques used (e.g., to de-escalate intense distress) were noted. In case of worsening of symptoms, the patient was referred to the emergency services in the hospital, and the referring clinical team was notified. All interventions were delivered by a single clinician (a trainee clinical psychologist) who was trained in diagnostic interviewing, neuropsychological assessment, and interventions, and supervised by a licensed, consultant clinical psychologist. Further details of the intervention are presented in Table 2.

**Table 2.** Elements of the Brief ICCT Pertaining to Thematic Components of Cognitive Retraining.

| **Thematic Component** | **Elements of the Intervention** | **Sessions In Which Included** |
| --- | --- | --- |
| Cognitive Stimulation | Connect-4, Mindful Colouring | 2, 3, 4 |
| Metacognitive Strategy Training | Education about cognitive processes, monitoring strategy use during tasks, and helping the patient to identify, modify, and review strategies | 1, 2, 3, 4, 5 |
| Generalisation | Discussions regarding daily routines and everyday occupational functions, Roleplay | 1, 2, 3, 4, 5 |

### Analysis

In the first phase of the study (adaptation of the intervention), survey data were qualitatively analyzed and tabulated to identify trends and themes. The data from the ‘try-out trial’ was also qualitatively analyzed to identify challenges and pitfalls.

Data from the second phase of the intervention were analyzed using IBM SPSS 28.00. Descriptive statistics, including mean, standard deviation, minimum, and maximum values, were used to summarize sociodemographic, clinical, and baseline cognitive data. The Shapiro-Wilk Test of Normality (α = 0.05) was used to assess data distribution. Intervention efficacy was analyzed using paired-samples t-tests for normally distributed data and the Wilcoxon Signed-Rank Test for non-normally distributed data. Effect sizes were measured using Cohen’s d (Cohen, 1988). Change scores (Blanchard & Schwartz, 1988) were computed for the primary outcome measures individually for each participant using the following formula: {[(Pre-Intervention Score – Post-Intervention Score) / Pre-Intervention Score] * 100}. The change score was also derived for the mean values of all measures using the following formula: {[(Mean of Pre-Intervention Scores – Mean of Post-Intervention Scores) / Mean of Pre-Intervention Scores] * 100}. Additionally, the predictors of improvement were analyzed via Pearson’s Rank Order Correlation, followed by multiple linear regression.

No additional analyzes were performed beyond those prespecified, and all planned statistical analyzes were completed. There were no missing data; therefore, no imputation procedures were required.

## Results

### Survey Findings

The online expert survey included seven individuals with lived experience of depression and six clinicians (three psychiatrists, three clinical psychologists) from a tertiary psychiatric institute in India. Separate online survey forms were created for individuals with lived experience of depression and clinicians to capture various elements of cognitive deficits in depression and the degree to which these are addressed in routine clinical examination and treatment.

Individuals with depression reported significant cognitive difficulties, including attention deficits, memory issues, and challenges with planning, which impacted daily functioning. They perceived greater impairment in everyday life compared to clinicians, who noted that routine treatment often missed key aspects like executive dysfunction. Both groups agreed that cognitive impairments hinder recovery, with psychotherapy seen as more effective than pharmacotherapy. There was high demand for a cognitive training intervention, with patients prioritizing concentration, planning, and memory, while clinicians focused on attention, memory, and planning. Both groups preferred brief intervention formats, highlighting the need for targeted cognitive training to address depression-related deficits and improve functional outcomes (Fig 2).

**Fig. 1.**
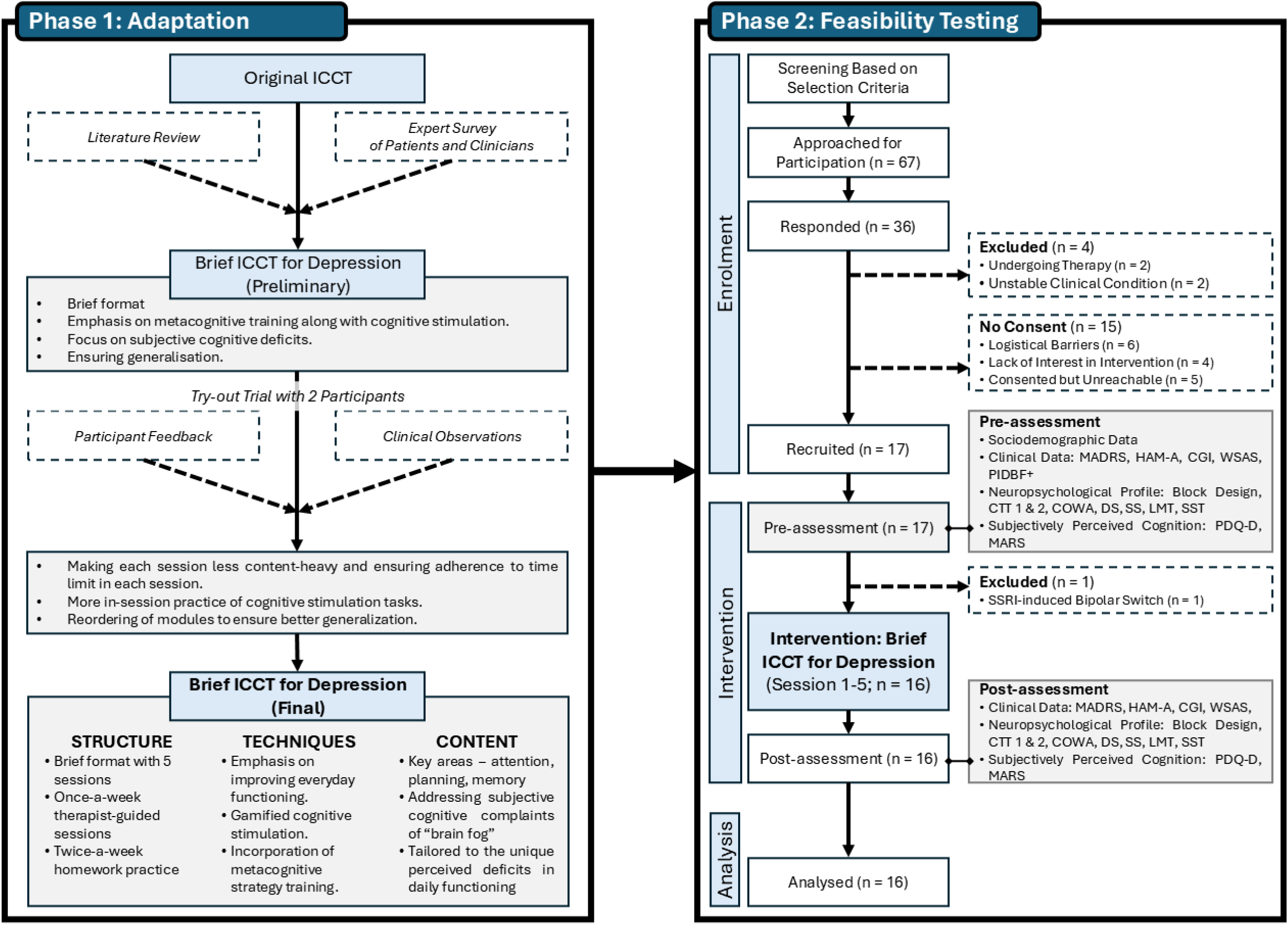
Study Flow

**Fig. 2.**
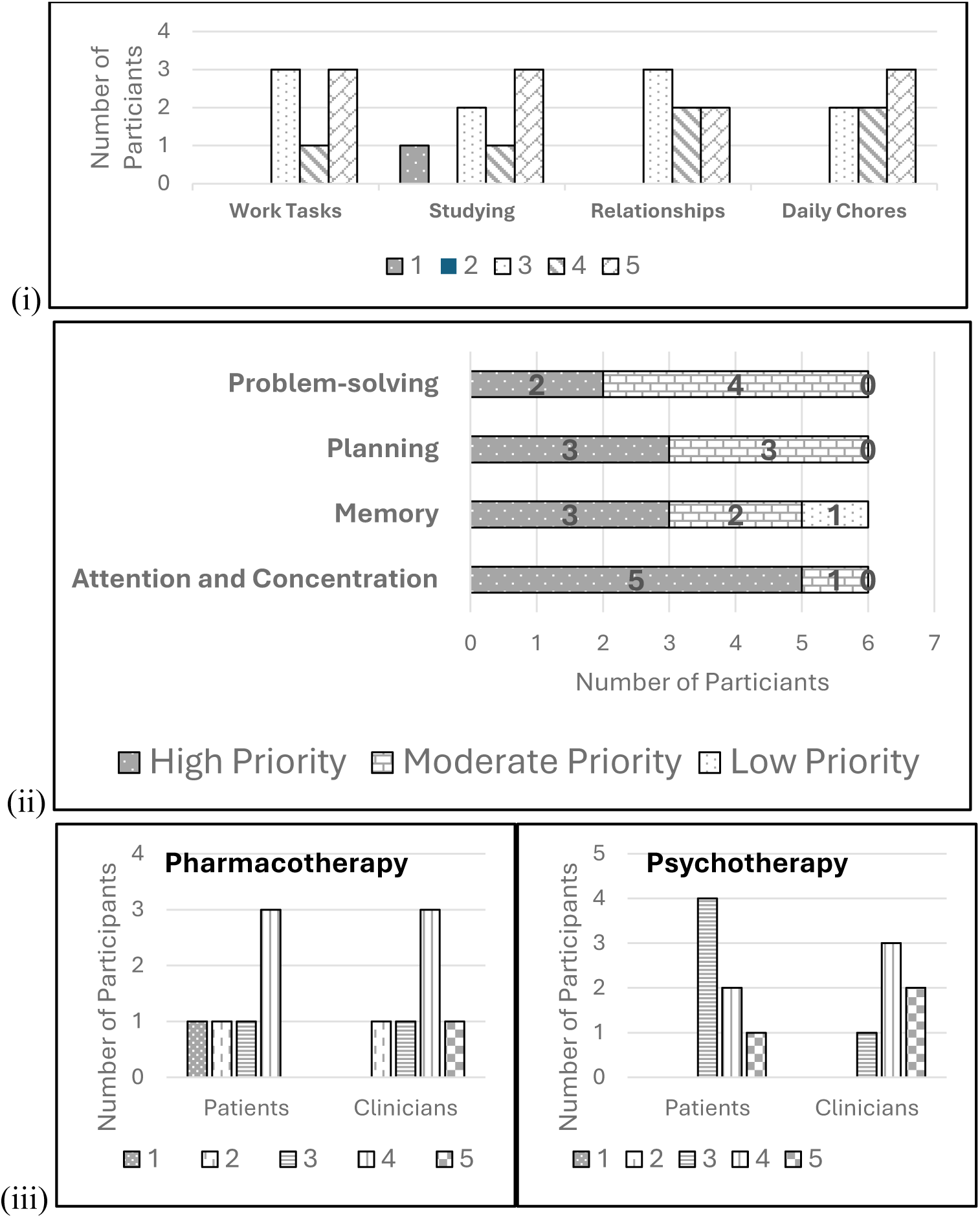
Diagrammatic Representation of Select Findings from Survey: (i) Ratings of Degree of Dysfunction (1 to 5, 5 being most dysfunction) in Various Domains Due to Cognitive Difficulties by Patients; (ii) Priority Ratings of Cognitive Domains as Intervention Targets by Patients; (iii) Perceptions of Impact of Treatment-as-Usual – Pharmacotherapy and Psychotherapy(rated on a scale of 1-5, 5 indicating greater effectiveness of treatment) on Cognitive Difficulties among Two Sample Groups

**Fig. 3.**
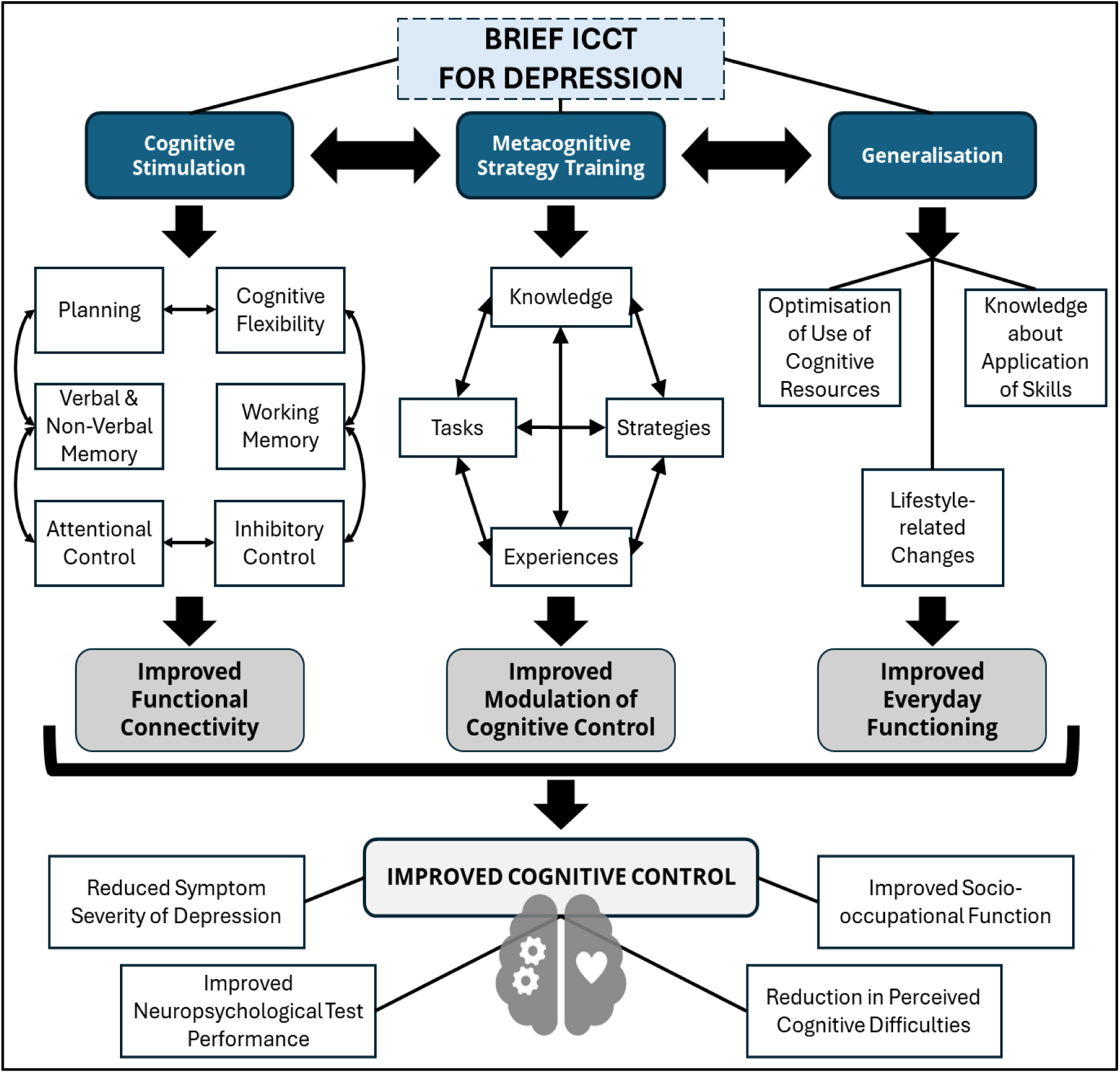
Proposed Mechanism of Action of the Brief ICCT for Depression

Hence, based on the aforementioned findings, the intervention was adapted to a brief format (3–5 sessions) with weekly therapist-guided sessions and homework practice. It emphasized everyday functioning, included metacognitive strategies and gamified cognitive stimulation, and focused on attention, planning, and memory, while addressing subjective complaints and individualized functional deficits.

### Sample Characteristics

The sample had an average age of 30.1 ± 4.75 years and predominantly included women (75%), professionals (62.5%), and individuals from middle- to upper-socioeconomic strata (81.2%). Most participants were well-educated (75% were graduates or higher). The sample description is provided in Table 3.

**Table 3.** Sociodemographic Characteristics (Categorical Variables) of Sample.

| Variable | Categories | Frequency (Percentage) |
| --- | --- | --- |
| Gender | Female | 12 (75.0) |
|  | Male | 4 (25.0) |
| Occupation | Homemaker | 2 (12.5) |
|  | Skilled | 1 (6.2) |
|  | Professional | 9 (62.5) |
|  | Student | 2 (12.5) |
|  | Unemployed | 2 (12.5) |
| Religion | Hindu | 14 (87.5) |
|  | Muslim | 1 (6.2) |
|  | Christian | 1 (6.2) |
| Socioeconomic Status | Lower | 3 (18.7) |
|  | Middle | 8 (50.0) |
|  | Upper | 5 (33.3) |
| Marital Status | Unmarried | 8 (50.0) |
|  | Married | 6 (37.5) |
|  | Divorced/Separated | 2 (12.5) |
| Education | High School | 4 (25.0) |
|  | Graduate | 9 (56.2) |
|  | Post-graduate | 3 (18.7) |

Characteristics of dropouts and individuals who declined participation were examined to assess potential recruitment bias and are reported in the supplementary material.

### Treatment Effects

A paired sample t-test was conducted to assess the intervention’s effectiveness for normally distributed data (Table 4), with Cohen’s d used to measure effect size (small: 0.2–0.5, medium: 0.5–0.8, large: >0.8). The results showed significant improvement (p < 0.05) with medium to large effect sizes across all variables. The highest effect sizes were observed in MADRS, HAM-A, DS Forward, the combined Digit Span and Spatial Span total, and PDQ-D. The Wilcoxon Signed Ranks Test was used to evaluate the intervention’s effectiveness for non-normal and categorical data. Results (Table 4) showed significant improvement (p < 0.05) in CGI scores, all PDQ-D domains (attention and concentration, retrospective memory, prospective memory, and planning and organization), and Go Omission errors on the SST. No significant improvement was found in other SST variables. Positive changes were seen in all outcomes, with the greatest improvements in GoMISS (SST), MADRS, HAM-A, LMT (Story Units), Retrospective Memory, and Planning/Organization (PDQ).

**Table 4.**
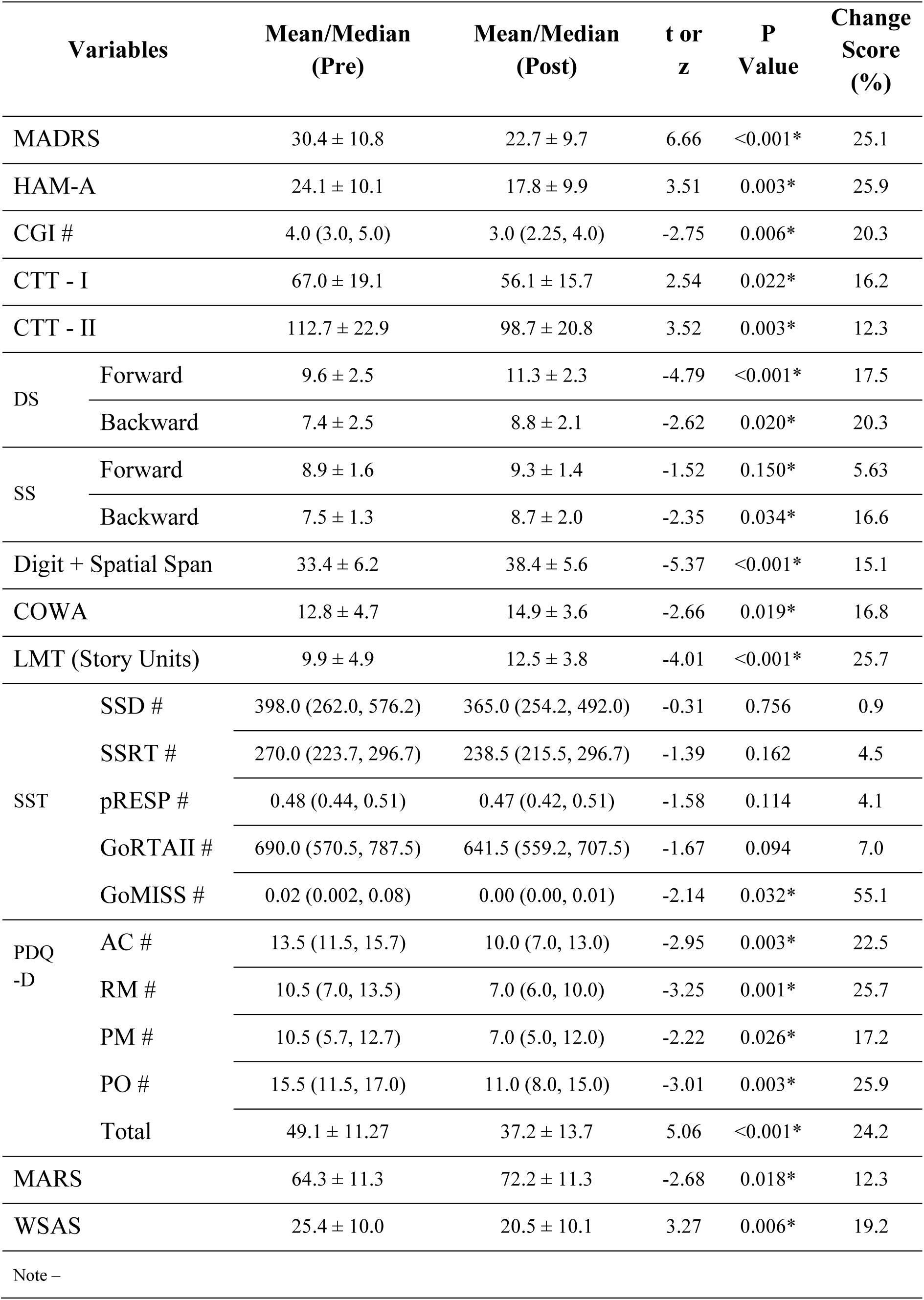
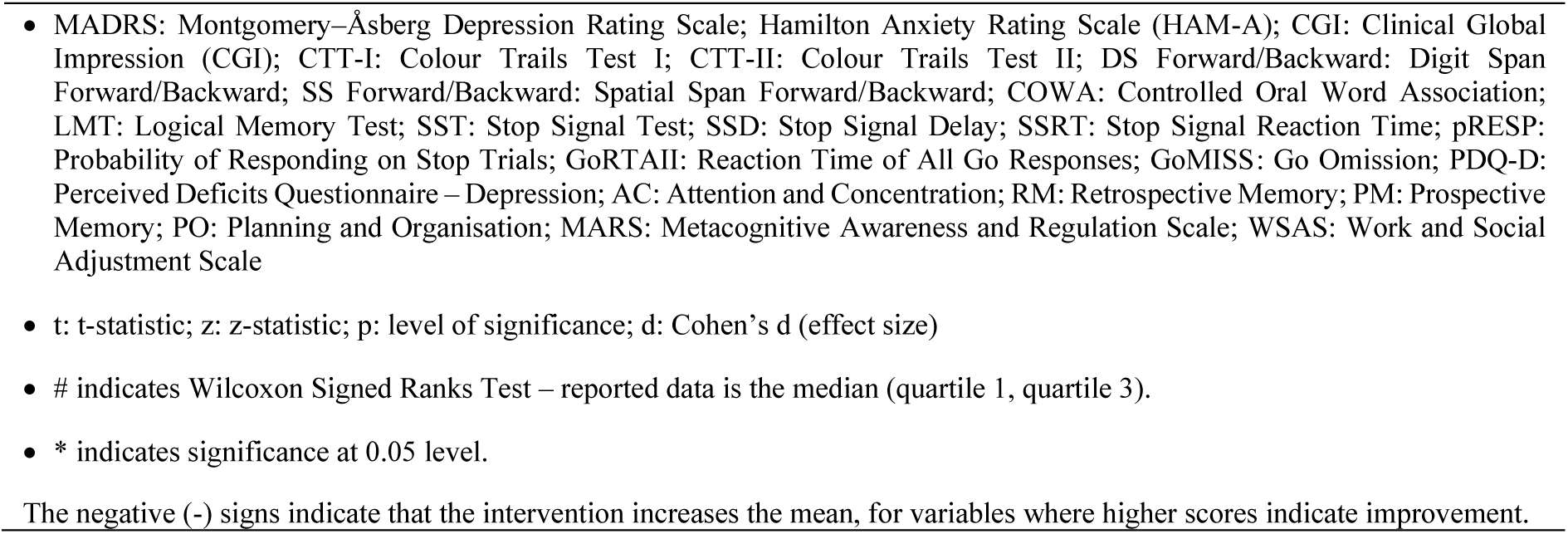
Paired Sample t-Tests and Wilcoxon Signed Ranks Test for Outcome Variables.

### Feasibility of Interventions

Feasibility was measured using a checklist developed by the researcher, based on the criteria for study feasibility outlined by Bowen and colleagues (2009). The checklist recorded data about the acceptability, demand, implementation, practicality, and integration of the intervention, as per Bowen’s criteria (Bowen et al., 2009).

High acceptability was evident from participant feedback, with ratings showing high satisfaction, perceived everyday changes, and a strong willingness to recommend the program. Module-specific preferences highlighted that the ‘planning and organization’ module had the highest engagement and perceived usefulness. Demand was reflected in the absence of dropouts among participants, with one participant excluded due to clinical reasons. Efforts such as flexible scheduling, reminders, and hybrid delivery modes (online and offline) likely contributed to retention. Implementation showed that 79% of sessions fully adhered to the manual, with minimal use of unrelated techniques to manage distress. Practicality was demonstrated by consistent attendance and homework completion despite logistical challenges, affirming the intervention’s feasibility in resource-constrained settings.

## Discussion

As a proof-of-concept investigation, this study aimed to adapt the ICCT intervention into a brief format suited for depression and to generate preliminary evidence of feasibility and efficacy. The intervention was adapted for use with a sample of individuals with depression. Following this, the intervention was administered on a limited sample of individuals with depression to test the feasibility and effectiveness of the intervention.

The Brief ICCT led to improvements across clinical status, neuropsychological performance, subjective cognitive complaints, and socio-occupational functioning. Significant gains were seen in attention, cognitive flexibility, working memory, verbal fluency, and memory, with the largest effects in attention, working memory, and learning. These improvements in cognitive control are especially relevant for depression. There is preliminary evidence to suggest that impairments in cognitive control (specifically, cognitive flexibility and working memory) precede the onset of depression, as demonstrated in a rat model (Maramis et al., 2021). Further, the role of cognitive control in emotion regulation, rumination, and relapse risk in depression is well established (Murphy et al., 2011; Beckwé et al., 2013; Figueroa et al., 2019; Kashyap et al., 2022). Hence, the improvement shown in cognitive control with minimal sessions, including untrained tasks and domains, is particularly relevant.

No significant improvements were seen in most SST variables, except for ‘GoMISS’, which reflects missed ‘Go’ responses. This may be due to the absence of specific response inhibition tasks in the intervention, even though other untrained cognitive functions showed significant gains. Prior studies on ICCT have shown small effects on response inhibition (Rini et al., 2019), and broader literature suggests this domain is generally resistant to change through cognitive training. A meta-analysis by Li et al. (2022) found that even intensive 30-day stop-signal and go/no-go training yielded limited improvements.

The current intervention emphasized addressing subjectively perceived cognitive deficits through metacognitive strategy training, resulting in significant improvements in PDQ-D and MARS scores. The highest improvement (25%) was in the PDQ-D planning and organization subdomain, aligning with participant feedback and clinician-rated engagement. Research shows interventions targeting executive functions, psychoeducation, lifestyle changes, and metacognition can alleviate perceived cognitive decline (Buitenweg et al., 2019; Goghari & Savage, 2018). The observed decrease in PDQ-D scores is meaningful, given its association with daily functioning and quality of life (Chokka et al., 2019; Zhang et al., 2020).

Despite these domains not being directly trained, clinician-rated depression, anxiety, and CGI severity scores improved significantly with high effect sizes. This finding supports the established link between mood symptoms and cognitive deficits (Harvey, 2011; Zimmerman et al., 2018) and echoes previous ICCT studies in OCD that showed reductions in MADRS and CGI scores (Rini et al., 2019; Bhattacharya, 2023). Although evidence for cognitive training in depression is mixed (Iacoviello et al., 2018; Elgamal et al., 2007; Morimoto et al., 2014; Banerjee, 2020), this study achieved robust improvements even with its shorter duration.

A central aspect of the brief ICCT was improving everyday functioning and restoring socio-occupational functioning. Results indicate that the Brief ICCT achieved significant improvements in WSAS scores, which are indicators of everyday functioning in the domains of work, relationships, and leisure activities and are related to depression severity, as per the STAR*D trial (IsHak et al., 2016). Cognitive deficits are closely linked to everyday functioning (Chokka et al., 2019; Zhang et al., 2020) and have been shown to mediate the relationship between depression and role functioning (Buist-Bouwman et al., 2008). Idiographic cognitive impairments also predict functional outcomes (Tran et al., 2021), and perceived cognitive deficits further contribute to this relationship. Consistently, the STAR*D trial found that depression severity correlates with socio-occupational functioning as measured by WSAS scores (IsHak et al., 2016). Hence, in the current study, the observed improvement in everyday functioning can be attributed to the intervention’s comprehensive approach, which targets cognitive functions (e.g., attention and concentration), perceived cognitive deficits, and depressive symptoms. Given that narrow transfer remains a major shortcoming of existing cognitive training interventions for depression (Woolf et al., 2021; Goldberg, Kuslak, & Kurtz, 2023), the current findings, demonstrating significant gains in functional outcomes, mark an important advancement in bridging this gap.

Feasibility was evaluated using Bowen et al.’s (2009) framework. The intervention demonstrated high acceptability – participants reported strong satisfaction, noticeable improvements in daily functioning, and a high willingness to recommend it to others. The most engaging component was Module 2 (“Planning and Organization”), likely due to its emphasis on planning and sleep – areas frequently disrupted in depression and linked to functional impairment (Sarıarslan et al., 2015; Murphy & Peterson, 2017; Kornilaki, 2021; Liang et al., 2023). Homework compliance was high (78%), consistent with cognitive training studies (49– 93%; Howe et al., 2018), and aligns with evidence from psychotherapy research showing that adherence improves outcomes (Mausbach et al., 2010; Kazantzis et al., 2016). Retention was strong, supported by hybrid and flexible scheduling. However, higher participation rates among younger and middle- to upper-SES individuals highlight the need to explore and address barriers to access for older adults and those from lower-income backgrounds.

The Brief ICCT incorporated the three pillars of cognitive training as outlined by Bowie (2016): cognitive stimulation, metacognitive strategy training, and generalization to everyday functioning. Cognitive stimulation may have contributed to the neuropsychological improvements observed in the current study, potentially by enhancing functional connectivity, a mechanism suggested by prior research (Eack et al., 2009; Baune & Renger, 2014; Chapman et al., 2015). While this remains a hypothesis in the current context, it is particularly relevant for depression, where brief interventions may more readily influence functional connectivity. Future studies are needed to directly examine these neural changes. Metacognitive strategy training likely enhanced both cognitive control and metacognitive awareness, consistent with Roebers’ (2017) view that these functions are interlinked and rooted in shared neurobiological mechanisms such as the prefrontal and anterior cingulate cortices. Generalization – the transfer of skills from therapy to daily life – was central to the intervention’s design (Best & Bowie, 2017). By combining cognitive control via cognitive stimulation and metacognitive strategies, the intervention may have facilitated far transfer effects, supporting broader functional improvements (Greenwood & Parasuraman, 2016).

### Strengths and Limitations

As a proof-of-concept study, the current investigation aimed to adapt the ICCT into a brief intervention for patients with depression and to examine its feasibility and preliminary efficacy in a real-world clinical setting. The study was designed to generate initial evidence regarding the intervention’s clinical promise, acceptability, and potential impact on cognitive and functional outcomes, rather than to establish definitive efficacy, thereby informing the rationale and design of subsequent randomized controlled trials.

This study addressed a key treatment gap by offering a brief, structured intervention targeting cognitive deficits in depression. It uniquely combined metacognitive strategy training with a focus on generalization and assessed both subjective and objective outcomes. The ecologically valid sample and multidimensional assessments are additional strengths. The study also used alternate forms of neuropsychological assessments to control for practice effects. However, the lack of a control group, the small sample size, and the absence of independent checks of treatment fidelity limit generalizability. Homework compliance was not objectively assessed, and subgroup analyzes were not possible. Despite these limitations, the study addresses a unique treatment gap by offering proof-of-concept for a brief intervention targeting objectively assessed as well as subjectively perceived cognitive deficits in depression. Therefore, further studies are warranted to validate and expand upon these findings.

## Conclusion

This proof-of-concept study adapted and evaluated the Brief ICCT for Depression, showing its potential to address both objectively assessed and subjectively perceived cognitive impairments. By combining cognitive stimulation, metacognitive strategies, and generalization, the intervention offers a promising supplement to standard depression treatments. The findings lay the ground for further research to establish its long-term efficacy and broader applicability. These results add to the growing literature on innovative, scalable interventions for depression and offer important directions for clinical practice and future research.

## Data Availability

All data produced in the present study are available upon reasonable request to the authors

## Acknowledgements

The authors thank Dr. Binu V. S. for his contributions to the study design and statistical analysis, and Mr. Kuldeep Sharma for assistance in reviewing preliminary statistical analysis. We also acknowledge Ms. Uma V., Ms. Srijita Gupta, Ms. Manaswita Sinha, Ms. Padmaja Sarangi, and Ms. Anju George for their assistance with participant recruitment.

## Statements and Declarations

### Ethical Considerations

Collection of data was initiated after receiving the due approval from the Department Ethics Subcommittee and the Institutional Ethics Board (Number: NIMH/DO/BEH.Sc.Div/2023-24), and registration with the Clinical Trials Registry – India (CTRI; Registration No.: CTRI/2023/09/057529). Participants provided written informed consent after being explained the study in a language they understood and were informed about its experimental nature and lack of guaranteed benefit. Confidentiality was ensured through coded data storage and restricted access, with participants free to withdraw at any time without consequence; any clinical risks identified were appropriately referred for management. Written informed consent to participate in the study was sought from all participants in the presence of a witness.

### Data Availability

Data will be made available on request.

