## Supplemental Material for "ADAPTATION AND FEASIBILITY OF A BRIEF, INTEGRATED COGNITIVE CONTROL TRAINING INTERVENTION FOR DEPRESSION: A PROOF-OF-CONCEPT TRIAL"

### Supplementary Material

#### S1. Analysis of Non-Consenting Participants

There were no dropouts among participants; one individual was excluded after two sessions due to a change in clinical status (SSRI-induced bipolar switch). Efforts such as flexible scheduling, reminders, and hybrid delivery may have contributed to high retention. However, when a closer look is taken at the proportion of individuals approached for participation (n = 67), those who responded (n = 36), and those who finally consented to participation (n = 17), the following trends emerged. The primary reasons for non-consent to participate were logistical constraints, including time limitations, scheduling difficulties, and competing personal or occupational commitments.

The ages of participants and non-participants (those who did not consent to the intervention) were also compared using the Mann-Whitney U Test. The results (z = 602.000, p = 0.011) indicated that the non-participants group had significantly higher ages (mean rank = 37.54 years) than the participants group (mean rank = 23.59). This indicates that, in general, younger individuals chose to participate in the program, and that demand for the program is higher among them. Chi-square tests comparing participants and non-participants showed that non-participants were predominantly from lower- and middle-socioeconomic strata, while participants were mainly from middle- and upper-socioeconomic strata.

Table S1. Chi-Square Tests Comparing Participants with Non-Participants

| Variables | | Participants (Frequency, %) | Non-Participants (Frequency, %) | $\chi 2$ | p |
|---|---|---|---|---|---|
| Sex | F | 13, 76.4% | 28, 56.0% | 2.239 | 0.135 |
| | M | 4, 23.5% | 22, 44.0% | | |
| Socioeconomic Status | LSES | 4, 23.5% | 25, 50.0% | 7.645 | 0.022 |
| | MSES | 6, 35.2% | 19, 38.0% | | |
| | USES | 7, 41.1% | 6, 12.0% | | |

These findings suggest that, while demand for the intervention was high, as seen from the low dropout rate, practical barriers influenced participation, underscoring considerations of feasibility in routine clinical settings.
